# Neuron loss in the brain starts in childhood, increases exponentially with age and is halted by GM-CSF treatment in Alzheimer’s disease

**DOI:** 10.1101/2024.07.14.24310223

**Authors:** Stefan H. Sillau, Christina Coughlan, Md. Mahiuddin Ahmed, Kavita Nair, Paula Araya, Matthew D. Galbraith, Brianne M. Bettcher, Joaquin M. Espinosa, Heidi J. Chial, Neill Epperson, Timothy D. Boyd, Huntington Potter

**Affiliations:** Department of Neurology and the University of Colorado Alzheimer’s and Cognition Center, University of Colorado Anschutz Medical Campus, Aurora, CO; Linda Crnic Institute for Down Syndrome, University of Colorado, Anschutz Medical Campus, Aurora, CO; Department of Clinical Pharmacy, Center for Pharmaceutical Outcomes Research (CePOR); Department of Pharmacology, University of Colorado Anschutz Medical Campus, Aurora, CO; Department of Psychiatry, University of Colorado Anschutz Medical Campus, Aurora, CO

**Keywords:** Aging, Neurodegeneration, Alzheimer, granulocyte-macrophage colony stimulating factor, ubiquitin C-terminal hydrolase-L1, neurofilament light, glial fibrillary acidic protein, senolysis, neuroprotection

## Abstract

Aging increases the risk of neurodegeneration, cognitive decline, and Alzheimer’s disease (AD). Currently no means exist to measure neuronal cell death during life or to prevent it. Here we show that cross-sectional measures of human plasma proteins released from dying/damaged neurons (ubiquitin C-terminal hydrolase-L1/UCH-L1 and neurofilament light/NfL) become exponentially higher from age 2-85; UCH-L1 rises faster in females. Glial fibrillary acidic protein (GFAP) concentrations, indicating astrogliosis/inflammation, increase exponentially after age 40. Treatment with human granulocyte-macrophage colony-stimulating factor (GM-CSF/sargramostim) halted neuronal cell death, as evidenced by reduced plasma UCH-L1 concentrations, in AD participants to levels equivalent to those of five-year-old healthy controls. The ability of GM-CSF treatment to reduce neuronal apoptosis was confirmed in a rat model of AD. These findings suggest that the exponential increase in neurodegeneration with age, accelerated by neuroinflammation, may underlie the contribution of aging to cognitive decline and AD and can be halted by GM-CSF/sargramostim treatment.

## Introduction

Increasing age is the greatest risk factor for ‘natural’ age-associated cognitive decline (AACD) and, especially in females, for developing Alzheimer’s disease (AD), but the mechanisms underlying this connection are unknown ^1–10^. Neuronal loss and brain atrophy accompany aging and AD and can reasonably be inferred to lead to cognitive deficits ^11–14^. Increased inflammation is also correlated with AD pathogenesis and aging (termed ‘inflammaging’), but whether inflammation causes and/or is a response to neurodegeneration is also unknown ^1,8,15–17^. For example, early studies of patients with the inflammatory disease rheumatoid arthritis (RA) found that they exhibited a reduced risk of developing AD, which was attributed to their use of non-steroidal anti-inflammatory drugs (NSAIDs). However, studies of other inflammatory diseases, such as periodontitis, detected an increase in AD risk, and NSAID treatment in clinical trials showed no benefit to participants with either AD or mild cognitive impairment (MCI) (for discussion see ^16,18,19^). Evidently, the role of inflammation in aging and neurodegenerative disease is complex.

The identification of biomarkers of brain aging and the underlying mechanism(s) that they reflect are essential to advancing the development and testing of therapies for AD and for the more complex AACD ^17,20–23^. Here we assessed two proteins that reflect neuronal loss (UCH-L1) and axonal damage (NfL) and find that their plasma concentrations become exponentially higher throughout life starting from age 2. Furthermore, astrogliosis, measured by plasma concentrations of GFAP, becomes exponentially higher starting at age 40 and is thus likely to be reactive to rather than causal of neurodegeneration during aging and reflect a positive feedback loop that underlies the exponential nature of the increases in all three biomarkers of brain damage with age. Finally, we report that GM-CSF/sargramostim, a long-approved drug for stimulating immune stem cells in patients with leukopenia, which we previously found to improve cognition and biomarkers of neurodegeneration in a clinical trial of mild-to-moderate Alzheimer’s disease, effectively reverses the rate of neuronal loss in trial participants to the very low levels observed in early childhood in the current study. We also found that GM-CSF treatment reduces neuronal apoptosis in the hippocampi of aged TgF344-AD rats, a model that recapitulates all human AD brain neuropathology. These findings in humans and animals add biological significance to the statistically significant finding in the clinical trial.

## Results

### Biomarkers of Neurodegeneration

To identify biomarkers of neuronal cell loss or damage during brain aging, we focused on UCH-L1 and NfL because of numerous reports that their plasma concentrations are well correlated with neuronal damage in, for example, neurodegenerative disease and traumatic brain injury (TBI) ^24–29^. Although UCH-L1 was initially discovered and named as a ubiquitin C-terminal hydrolase, it is likely that this is not its primary function, especially in neurons, as cells and animals lacking UCH-L1 are not defective in the ubiquitin proteosome system ^25,30^. Indeed, UCH-L1 is primarily a brain protein, making up 1-5% of the total protein in neurons, with some expression in endocrine tissue ^25,26^. Suggestions for the neuronal function of UCH-L1 include the regulation of energy metabolism and mitochondrial fusion, antioxidant activity, and synaptic activity ^26,30,31^. Regardless of its function, an increased concentration of UCH-L1 in the plasma is a sensitive and rapidly-responding measure of acute neuronal cell damage/degeneration after TBI, and UCH-L1 expression is greatly reduced in the AD brain, possibly due to loss of neurons or neuronal activity ^24–27^. NfL is an intermediate filament protein in neurons and an essential component of axons. Traumatic or disease-associated damage to axon tracks leads to the release of NfL into the cerebrospinal fluid (CSF) and ultimately into the plasma where it can be detected as a biomarker of axonal damage in numerous neurodegenerative diseases as well as in TBI ^25,28,32^.

### Plasma concentrations of UCH-L1 are exponentially higher with age from early childhood

Plasma concentrations of UCH-L1, a measure of neuronal cell loss, were assessed in 317 healthy control participants between age 2 and 85 from three observational studies. These included 103 healthy control participants from the Human Trisome Project (HTP) of the Linda Crnic Institute for Down Syndrome (i.e., healthy controls without Down syndrome (DS)), 69 healthy control participants from the University of Colorado Alzheimer’s and Cognition Center (CUACC) study of the role of inflammation in AD (termed Bio-AD) ^17^, and 145 healthy control participants from the multiple sclerosis (MS) biomarker study (see demographics in Methods). The assessments were determined using the Quanterix SIMOA^®^ platform, which was also used previously to assess plasma biomarkers in the “sargramostim/GM-CSF AD trial” in participants with mild-to-moderate AD ^33^. As shown in **Figure 1A**, plasma UCH-L1 concentrations increase exponentially with age across the entire age spectrum, from an estimated 6.22 pg/ml at age 2 to approximately 15.56 pg/ml at age 85 (estimated change per year = 1.110%, 95% CI: (0.716%, 1.505%), p = 5.504 * 10^(−8)). Graphical inspection of the data, and comparison to a spline fit (**Supplementary Figure 1A**), indicate that a log-linear relationship of UCH-L1 concentrations with age is an excellent fit (Pearson correlation estimate (replicates log averaged) = 0.30, 95% CI: (0.20, 0.40)), which is equivalent to an exponential relationship on the original scale (**Figure 1A and 1B**). Interestingly, most of the age-associated increase in plasma concentrations of UCH-L1 occurs in females (**Figure 1C**, estimated female change per year = 1.448%, 95% CI: (0.942%, 1.957%), p = 3.635 * 10^(−8); estimated male change per year = 0.582%, 95% CI: (−0.036%, 1.203%), p=0.0650; sex difference p=0.0342; estimate of the ratio of ratios = 0.99146, 95% CI: (0.98362, 0.99936)). The log plots in **Figure 1D** and the spline analyses (**Supplementary Figure 1B**) support the conclusion that the plasma concentration of UCH-L1 increases exponentially with age in males and females. Because plasma UCH-L1 is primarily derived from damaged brain neurons, its concentration in plasma provides a minimal estimate of the ongoing neuronal cell death in the brain with age (**Supplementary Figure 2**).

**Figure 1.**
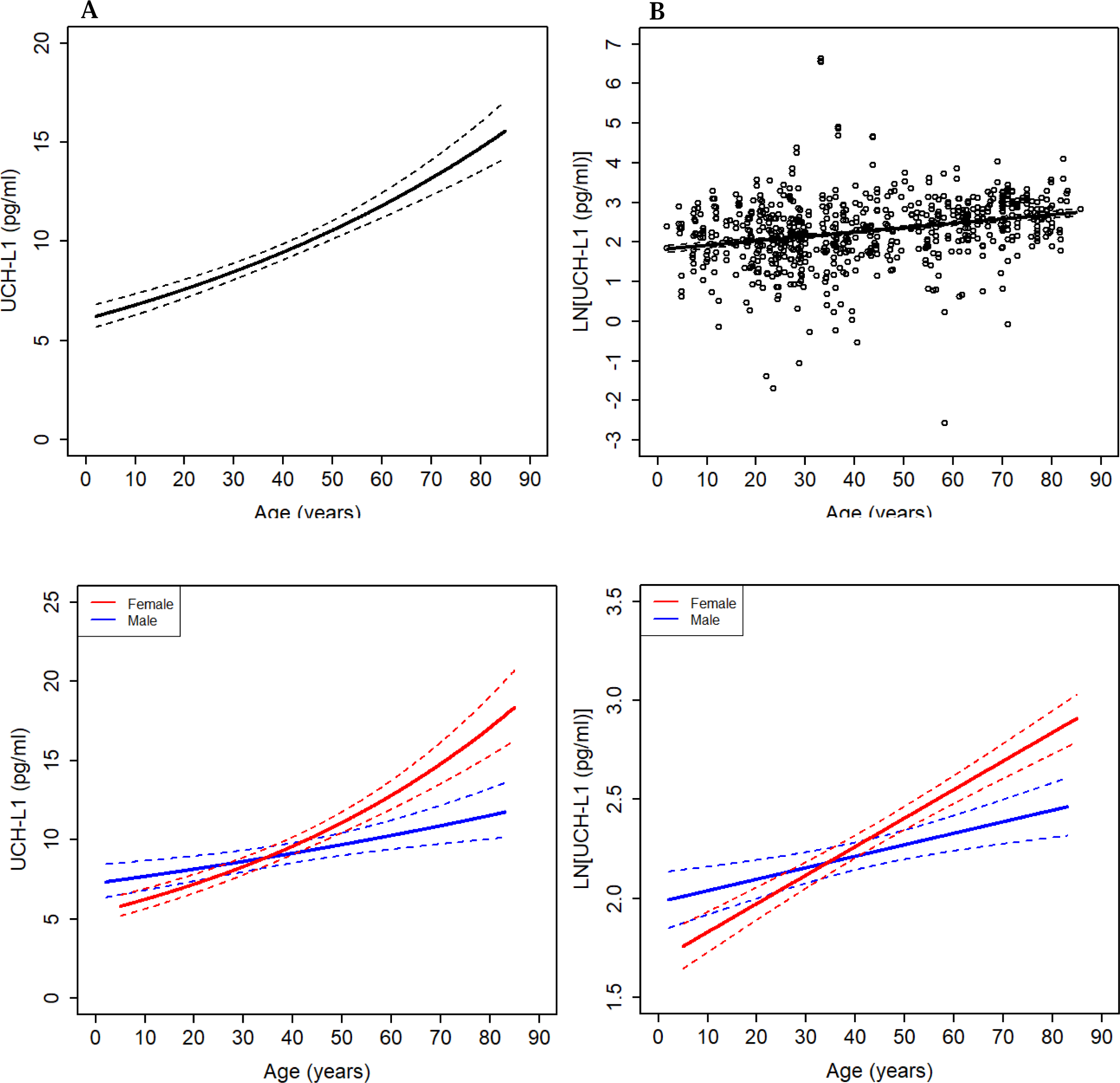
Plasma UCH-L1 concentrations are higher with advancing age in healthy control participants, especially in females. Concentrations of UCH-L1 in plasma from 314 healthy control participants who were part of either the Crnic Institute Human Trisome Project (n=103), the CUACC Bio-AD longitudinal observational study (n=69), or the MS healthy controls biomarker study (n=145) (3 participants lacked usable UCH-L1 data) were compared to the age of the participant. Together, these three healthy control cohorts span ages 2-85. The association between absolute UCH-L1 concentrations and age and the point wise standard errors are shown in (**A**). The UCH-L1 log plot with point wise standard errors is shown in (**B**). The data show that plasma UCH-L1 levels are exponentially higher with age across the lifespan (estimated change per year=1.110%, 95% CI: (0.716%, 1.505%), p = 5.504 * 10^(−8)). The effect of sex on the curve is shown in (**C**), which indicates that the majority of the promotion effect of age on plasma UCH-L1 concentrations are driven by females (estimated female change per year=1.448%, 95% CI: (0.942%, 1.957%), p = 3.635 * 10^(−8); estimated male change per year=0.582%, 95% CI: (−0.036%, 1.203%), p=0.0650; sex difference p=0.0342; sex ratio of ratios = 0.99146, 95% CI: (0.98362, 0.99936), p=0.0342). The effect of sex on the UCH-L1 log plot with point wise standard errors is shown in (**D**). The associations between absolute UCH-L1 concentrations and age and the point wise standard errors are shown. For the age range of 40-83 years, the area under the curve (AUC) average for the expected UCH-L1 values in males was statistically significantly less than the expected values in females (ratio estimate = 0.792, 95% CI: (0.628, 0.997), p value = 0.0472). Because the age effects in the model are linear in the log plot, the AUC average is equivalent to comparing the expected value for males to females at the midpoint of the range, 61.5 years.

### Plasma concentrations of NfL are exponentially higher with age from early childhood

Plasma concentrations of NfL, which is released primarily from damaged axons, is commonly used to assess/stage AD, MCI, and the risk of future cognitive decline ^22,28^. Therefore, we examined the effects of age and sex on plasma concentrations of NfL in the 317 healthy control participants. As shown in **Figure 2A** and **2B**, plasma concentrations of NfL were exponentially higher with increasing age in healthy control participants (p < 2.220 *10^(−16)), with the slope of the log-transformed curve being greater than that observed for UCH-L1 with increasing age, such that NfL plasma concentrations are higher by 2.469% per cross-sectional year (95% CI: (2.225%, 2.714%), p < 2.220 * 10^(−16)). Of note, the 95% confidence intervals for the NfL and UCH-L1 rates of increase are non-overlapping, suggesting a statistically significant difference (alpha=0.05) between the two. The exponential curve of plasma NfL with age showed a trend of being steeper for males than for females (p = 0.0502), with an estimated difference per year of 2.775% (95% CI: (2.388%, 3.164%), p < 2.220 * 10^(−16)) for males, compared to an estimated difference per year of 2.277% (95% CI: (1.965%, 2.590%), p < 2.220 * 10^(−16)) for females (Figure 2C); estimate of ratio of ratios = 1.00487, 95% CI: (1.00000, 1.00976), p=0.0502.

**Figure 2.**
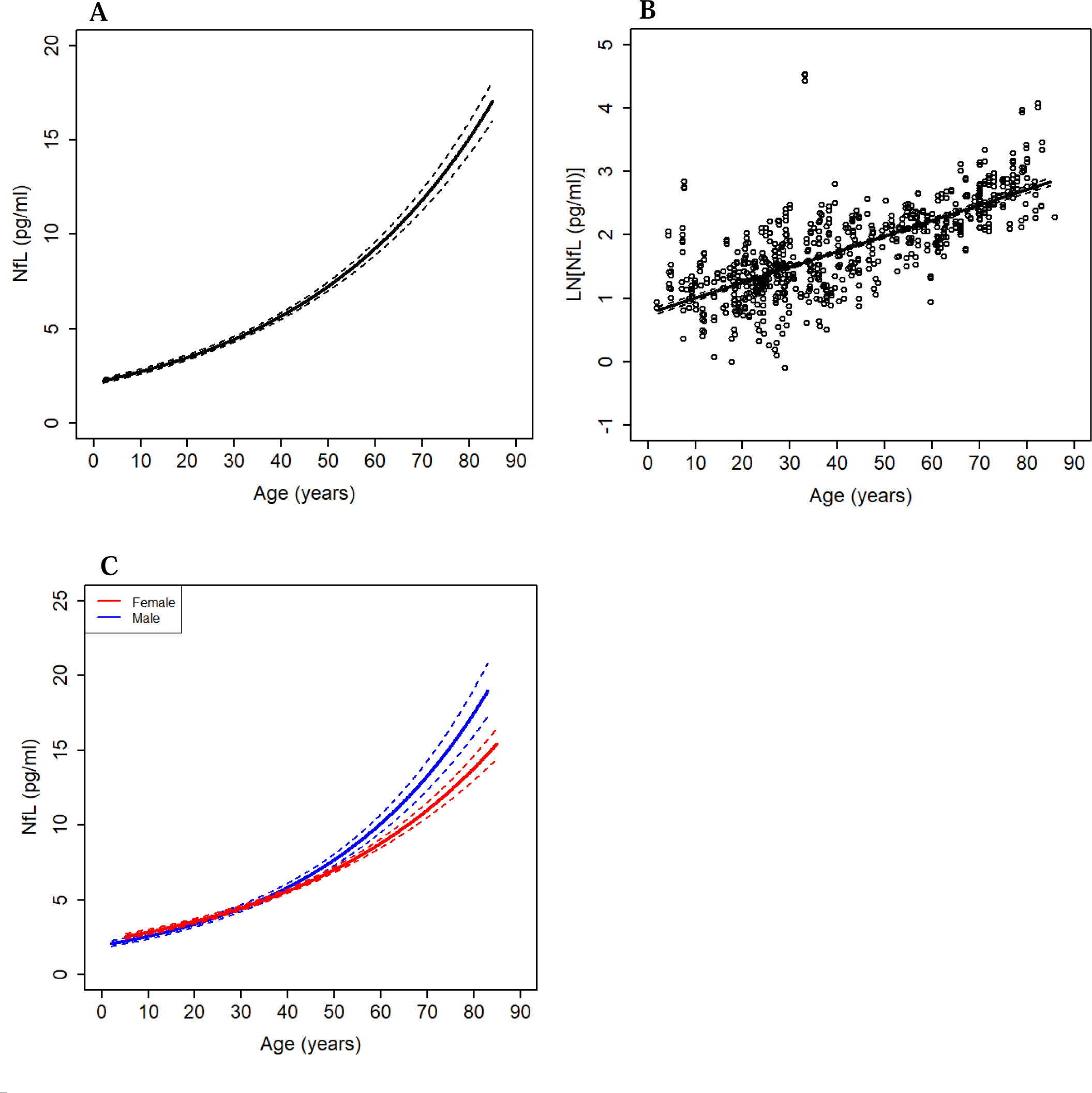
Plasma concentrations of NfL are exponentially higher with age in healthy control participants. Concentrations of NfL in plasma samples from the 317 healthy control participants who were part of the Human Trisome Project (HTP) (n=103), the Bio-AD longitudinal observational study (n=69), or the MS healthy controls biomarker study (n=145) were compared to the age and sex of the donor. The data show that plasma concentrations of NfL are exponentially higher with age across the entire lifespan (p < 2.220 *10^(−16)). The association between absolute NfL concentrations and age and the point wise standard errors are shown in (**A**) and the log plot is shown in (**B**) (estimated change per year=2.469% per year, 95% CI: (2.225%, 2.714%), p < 2.220 *10^(−16)). Separate plots for plasma concentrations of NfL in males and females are shown in (**C**). For NfL, the exponential rate of change determined cross-sectionally was marginally statistically non-significantly greater in males than in females (ratio of ratios = 1.00487, 95% CI: (1.00000, 1.00976), p = 0.0502); estimated change per year was 2.775% (95% CI: (2.388%, 3.164%), p < 2.220 *10^(−16)) in males, compared to 2.277% per year (95% CI: (1.965%, 2.590%), p < 2.220 *10^(−16)) in females. When comparing the log linear fit to splines for the association of NfL with age and sex in healthy controls (replicates averaged), the graph suggests that the rate of increase accelerates somewhat with older age, but a linear relationship is still an excellent approximation for all healthy controls (**Supplementary Figure 1C**) and when males and females are examined separately (**Supplementary Figure 1D**).

### Plasma concentrations of GFAP are exponentially higher from age 40

GFAP is upregulated in activated astrocytes, and its increased concentration in CSF or plasma is a marker of reactive gliosis in aging, neurodegenerative disease, and TBI ^17,27,34^. Both females and males in our combined three cohorts that span ages 2-85 showed an apparent overall exponential age effect for plasma GFAP concentrations (**Figure 3A, 3B, and 3C**) for both sexes (estimated female change per year=1.803%, 95% CI: (1.431%, 2.177%), p < 2.220 * 10^(−16); estimated male change per year=1.496%, 95% CI: (1.037%, 1.957%), p = 4.240 * 10^(−10)), with no significant difference between males and females. Interestingly, spline modeling of the plasma levels of GFAP (replicates averaged) show a deviation from linearity in the log plots in that the GFAP levels show a U shape with a decline from age 2 to 25, a constant rate, and then an exponential rise only starting from approximately age 40 (**Figure 3D**). The U shape spline plot is apparent for both sexes with females apparently rising slightly faster with age (**Figure 3E**). These findings indicate that brain astrogliosis/neuroinflammation follows and thus is likely a reaction to the earlier and ongoing age-associated neurodegeneration.

**Figure 3.**
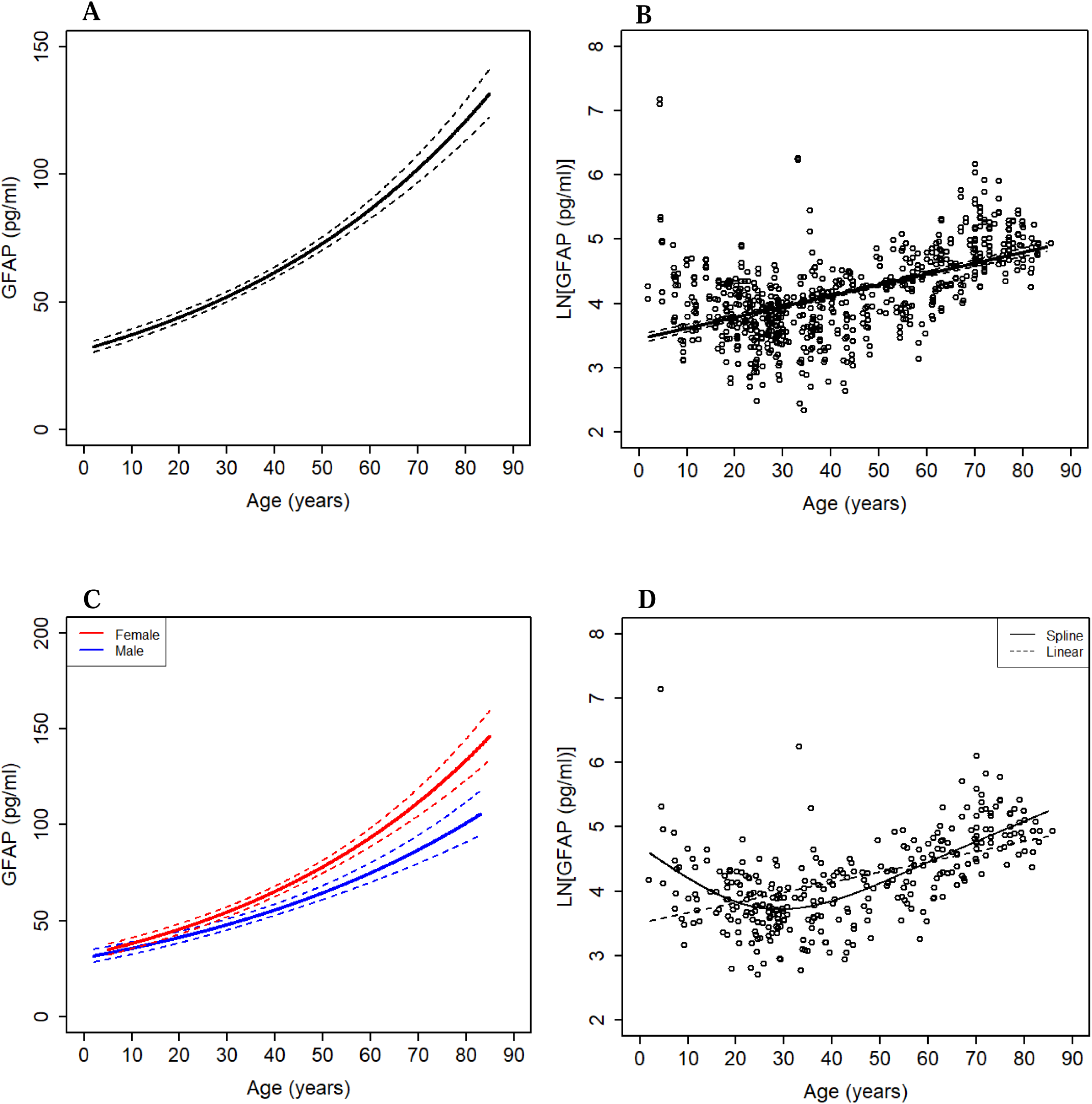
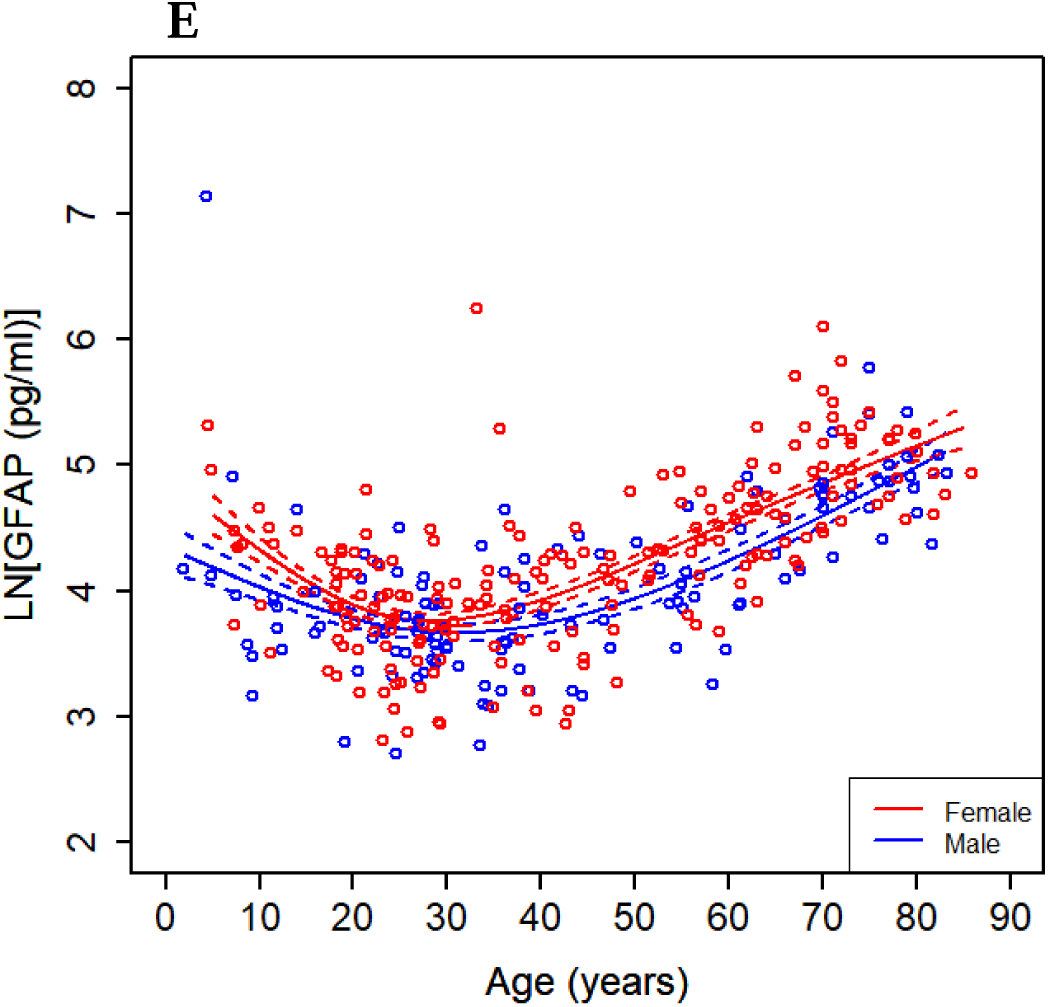
Plasma concentrations of GFAP are exponentially higher with age in healthy control participants. Concentrations of GFAP in plasma samples from the 317 healthy control participants who were part of the Human Trisome Project (HTP) (n=103), the Bio-AD longitudinal observational study (n=69), or the MS healthy controls biomarker study (n=145) were compared to the age and sex of the donor. The data show that plasma concentrations of GFAP are exponentially higher with age across the entire lifespan (p < 2.220 *10^(−16)). The association between absolute GFAP concentrations and age and the point wise standard errors are shown in (**A**) and the log plot is shown in (**B**) (estimated change per year=1.696%, 95% CI: (1.404%, 1.989%), p < 2.220 *10^(−16)). Separate plots for plasma concentrations of GFAP in males and females are shown in (**C**). Both sexes showed an exponential age effect for GFAP (estimated female change per year=1.803%, 95% CI: (1.431%, 2.177%), p < 2.220 *10^(−16); estimated male change per year=1.496%, 95% CI: (1.037%, 1.957%), p = 4.240 * 10^(−10)), but there was not a statistically significant difference in age effect between the sexes (ratio of ratios = 1.00302, 95% CI: (0.99720, 1.00889), p=0.3088). We also compared the log linear fit to splines for the biomarker association with age in the healthy controls (replicates averaged) in (**D**). The deviance test for comparing spline model to the null hypothesis of linear: p value < 2.2 * 10^(−16). The evidence is against linearity with a large sample, with the graph suggesting something of a U shape with GFAP concentrations accelerating significantly after age 40. A spline plot by sex in the healthy controls (replicates averaged) also suggests a U shape with GFAP concentrations accelerating significantly after age 40 in (**E**). Deviation from linearity is evidenced in the data from males and females, with females apparently rising slightly faster with age.

### Correlation analyses of plasma measures of age-associated brain degeneration

**Figure 4** shows the correlations of the measures of neurodegeneration (UCH-L1, NfL) and astrogliosis (GFAP) for all healthy control participants from age 2 to 85. All three measures (UCH-L1:NfL, UCH-L1:GFAP, and NfL:GFAP) are highly correlated with each other (P< 10^(−11)), indicating that brain aging across the lifespan reflects parallel increases in neurodegeneration, both cellular and axonal, and astrogliosis/inflammation.

**Figure 4:**
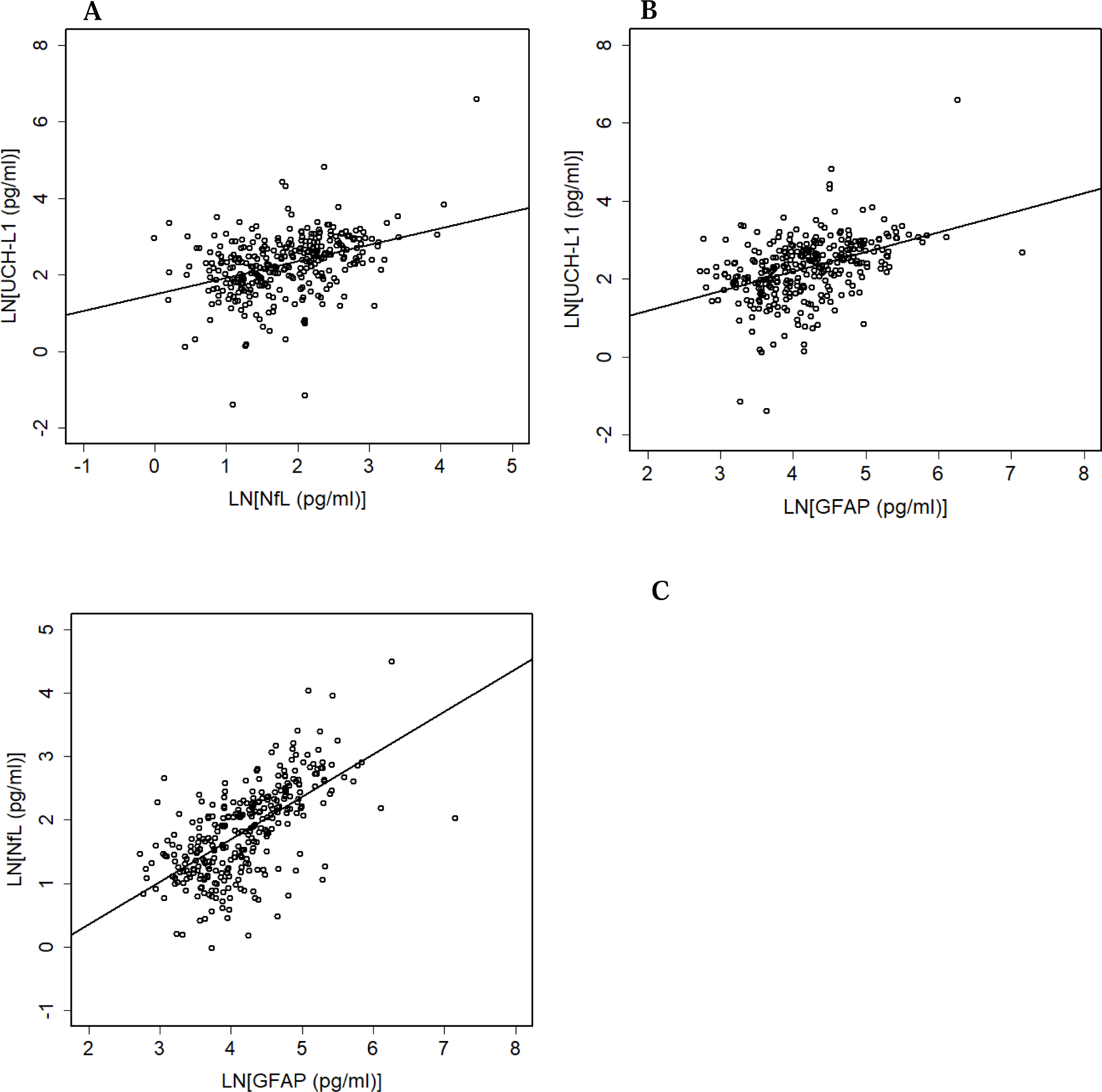
Correlations between plasma biomarkers of neurodegeneration and astrogliosis, UCH-L1, NfL, and GFAP. Log transformed replicate measures of UCH-L1, NfL, and GFAP from all normal control participants, age 2-85, were averaged over replicates and subjected to Pearson and Spearman correlation analyses and found to be highly correlated with each other. (**A**) UCH-L1:NfL, PC=0.37951, 95% CI: (0.280616, 0.470436), P = 3.157 * 10^(−12); (**B**) UCH-L1:GFAP, PC=0.42263, 95% CI: (0.327250, 0.509477), P = 4.441 * 10^(−15); and (**C**) NfL:GFAP PC=0.63743, 95% CI: (0.567089, 0.698536), P < 2.220 * 10^(−16).

### Comparisons of plasma measures of age-associated brain neurodegeneration and inflammation in MCI due to AD, mild-to-moderate AD, and healthy controls

AD dementia and its precursor, MCI due to AD, are both strongly associated with age and are accompanied by neurodegeneration and astrogliosis in the brain ^17^. Having established full age curves for plasma markers of neurodegeneration (UCH-L1 and NfL) and astrogliosis/inflammation (GFAP), we compared these to the age-associated concentrations of NfL, GFAP, and UCH-L1 in plasma samples from the 32 participants with MCI due to AD from the Bio-AD study (MCI) and in plasma samples from 36 participants with mild-to-moderate AD from our previously published GM-CSF/sargramostim clinical trial at baseline (Baseline AD GM-CSF Study Pooled) ^33^ (**Figure 5**). A diagnosis of MCI or mild-to-moderate AD was associated with higher overall levels of both NfL (**Figure 5A**) and GFAP (**Figure 5B**) compared to age-matched healthy control participants. NfL concentrations were higher in participants with mild-to-moderate AD than in participants with MCI, while there was no significant difference in GFAP concentrations between participants with mild-to-moderate AD and participants with MCI at 67.8 years, which is the mean age for the sargramostim/GM-CSF-treated mild-to-moderate AD participants (AD/HC estimate age 67.8: NfL: ratio estimate = 1.852, 95% CI: (1.514, 2.265), p = 2.201 * 10^(−7); GFAP: ratio estimate = 1.842, 95% CI: (1.539, 2.203), p = 4.169 * 10^(−9); MCI/HC estimate age 67.8: NfL: ratio estimate = 1.362, 95% CI: (1.104, 1.682), p=0.0050; GFAP: ratio estimate = 1.948, 95% CI: (1.473, 2.577), p = 2.230 * 10^(−5); AD/MCI estimate age 67.8: NfL: ratio estimate = 1.359, 95% CI: (1.042, 1.772), p=0.0242; GFAP: ratio estimate = 0.945, 95% CI: (0.699, 1.278), p=0.7091), as expected from previous studies ^17,28^. Interestingly, the plasma concentration of NfL showed age-associated higher levels in the participants with mild-to-moderate AD (2.714% per year, 95% CI: (0.625%, 4.847%), p=0.0122) (**Figure 5A**), whereas GFAP plasma concentrations did not show age-associated higher levels in the participants with mild-to-moderate AD (−0.030% per year, 95% CI: (−2.061%, 2.042%), p=0.9761). Plasma concentrations of both NfL and GFAP showed age-associated higher levels for MCI (NfL: 4.138% per year, 95% CI: (1.664%, 6.672%), p=0.0017; GFAP: 3.466% per year, 95% CI: (0.170%, 6.871%), p=0.0398) (**Figure 5B**). There were no statistically significant differences among the age slopes for healthy controls, AD, and MCI for either NfL or GFAP.

**Figure 5.**
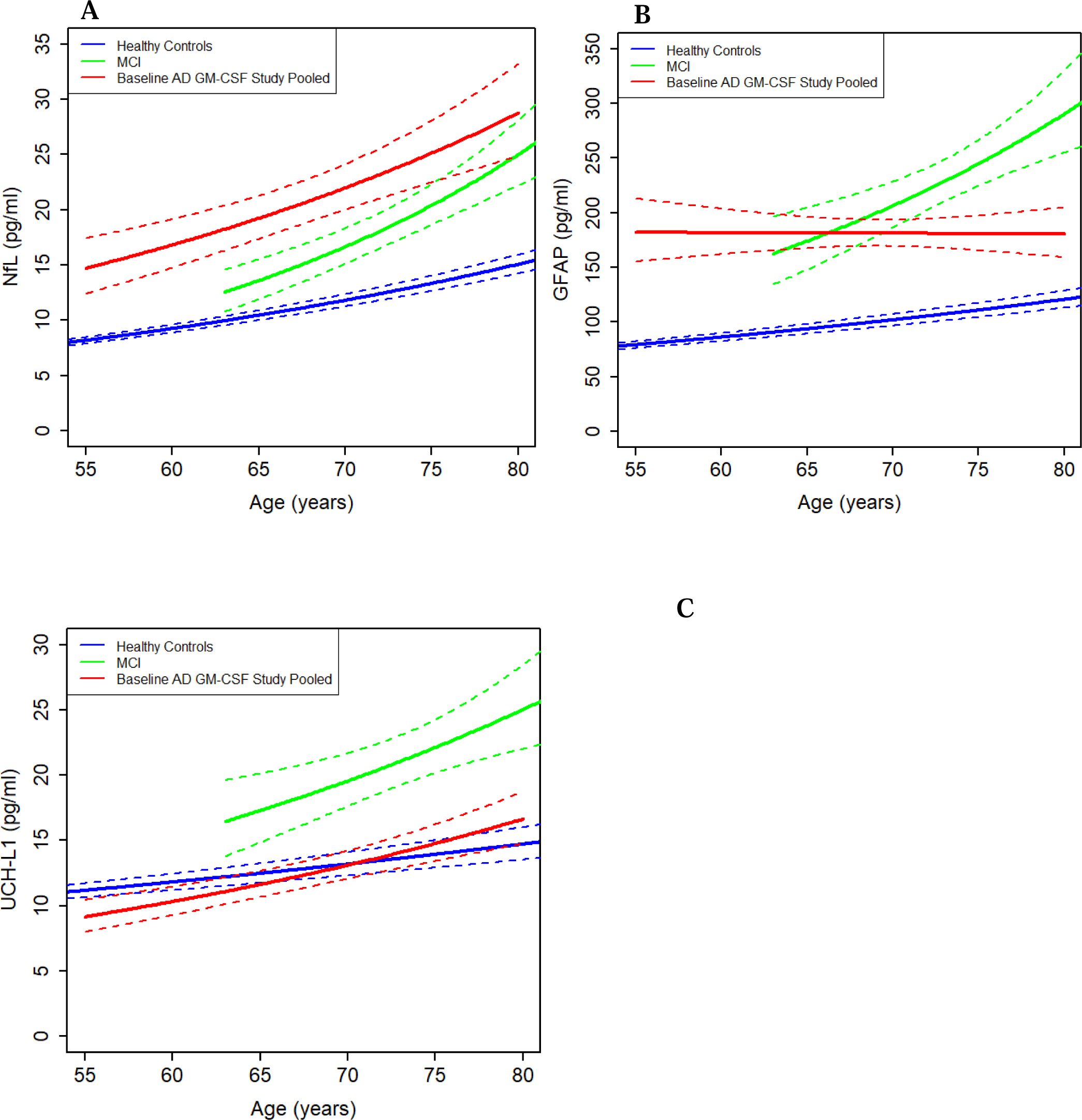
Comparisons between plasma NfL, GFAP, and UCH-L1 concentrations and age in participants with MCI due to AD, participants with mild-to-moderate AD, and healthy control participants. We compared the concentrations of NfL, GFAP, and UCH-L1 in plasma samples from participants assessed as having MCI due to AD as part of the CUACC Bio-AD longitudinal observational study (n=32) (MCI), and 36 participants from the Phase II, double-blind, randomized, placebo-controlled trial with recombinant human GM-CSF (the “sargramostim/GM-CSF AD trial”) at baseline prior to treatment with sargramostim/GM-CSF or placebo (Baseline AD GM-CSF Study Pooled), and the corresponding segment of the age curve from the 317 healthy control participants who were part of either the Crnic Institute Human Trisome Project (HTP) (n=103), the CUACC Bio-AD longitudinal observational study (n=69), or the MS healthy controls biomarker study (n=145). The mean age for the sargramostim/GM-CSF-treated mild-to-moderate AD participants was 67.8 years. (**A**) Plasma NfL concentrations in participants with MCI or mild-to-moderate AD are higher, correlating with disease progression, than in age-matched healthy control participants and also show an increase with age (MCI geometric mean estimate 15.27 pg/ml (95% CI: (12.54, 18.58) pg/ml) at age 67.8; mild-to-moderate AD geometric mean estimate of 20.75 pg/ml (95% CI: (17.22, 25.01) pg/ml) at age 67.8 and baseline calibrated compared to age-matched healthy control participants (geometric mean estimate of 11.21 pg/ml (95% CI: 10.36, 12.13) pg/ml; MCI/HC ratio estimate = 1.362, 95% CI: (1.104, 1.682), p=0.0050; AD/HC ratio estimate = 1.852, 95% CI: (1.514, 2.265), p = 2.201 * 10^(−7)). NfL was statistically significantly higher in AD than in MCI at 67.8 years (ratio estimate = 1.359, 95% CI: (1.042, 1.772), p=0.0242). The estimated exponential change in plasma NfL for MCI was 4.138% per year (95% CI: (1.664%, 6.672%), p=0.0017) and for mild-to-moderate AD was 2.714% per year (95% CI: (0.625%, 4.847%), p = 0.0122). The differences between the MCI or mild-to-moderate AD rate of change and healthy control rate of change were not statistically significant for NfL, but the power of the comparison test is limited because of the small sample size of the mild-to-moderate AD group (interaction test p value = 0.3953). (**B**) Participants with MCI or mild-to-moderate AD exhibit a higher concentration of plasma GFAP (MCI: geometric mean estimate = 191.78 pg/ml, 95% CI: (147.18, 249.89) pg/ml) (AD: geometric mean estimate of 181.28 pg/ml (95% CI: (155.40, 211.46) pg/ml) at the sargramostim/GM-CSF mean age of 67.8 and baseline calibrated) compared to age-matched healthy control participants (geometric mean estimate of 98.43 pg/ml (95% CI: (89.51, 108.24) pg/mL; MCI/HC ratio estimate = 1.948, 95% CI: (1.473, 2.577), p = 2.230 * 10^(−5); AD/HC ratio estimate = 1.842, 95% CI: (1.539, 2.203), p = 4.169 * 10^(−9)). GFAP levels for mild-to-moderate AD and MCI were similar at 67.8 years (AD/MCI ratio estimate = 0.945, 95% CI: (0.699, 1.278), p=0.7091). The levels for the MCI participants increase with age (estimated change per year = 3.466%, 95% CI: (0.170%, 6.871%), p=0.0398), but the levels do not change for the mild-to-moderate AD participants (estimated change per year = −0.030%, 95% CI: (−2.061%, 2.042%), p = 0.9761). The differences between the MCI or AD rate of change and healthy control rate of change were not statistically significant for GFAP, but the power of the comparison test is limited because of the small sample size of the mild-to-moderate AD group (interaction test p value = 0.1482). (**C**) The mean of plasma UCH-L1 concentrations in participants with MCI due to AD is statistically significantly higher (ratio estimate = 1.440, 95% CI: (1.101, 1.885), p=0.0088, at mean age of 67.8 years) than for healthy control participants, indicating the initiation of neurodegeneration beyond that caused by normal aging alone. Notably, the mean plasma concentrations of UCH-L1 at baseline for mild-to-moderate AD participants in the sargramostim/GM-CSF AD trial are similar to the concentrations in the healthy control participants at the corresponding age (ratio estimate = 0.967, 95%: (0.798, 1.171), p = 0.7257), possibly because neuronal loss has reduced the numbers of neurons available to release UCH-L1, but the increase with age remains statistically significant (estimated change per year = 2.420%, 95% CI: (0.783%, 4.083%), p=0.0054). The estimated slopes of the association between plasma UCH-L1 concentrations and age in the participants with MCI due to AD (estimated change per year=2.499%, 95% CI: (−0.446%, 5.531%), p = 0.0941) or with mild-to-moderate AD are between 2 to 2.5, which is the slope of the correlation between plasma UCH-L1 concentrations and age in the healthy control participants in **Figure 1** (estimated change per year=1.110%, 95% CI: (0.716%, 1.505%), p = 5.504 * 10^(−8)), but the differences are not statistically significant, most likely due to the small numbers of participants in the MCI due to AD group and in the mild-to-moderate AD group (interaction test p value = 0.2040).

UCH-L1 levels in the participants with MCI due to AD are higher overall than the plasma UCH-L1 concentrations of the healthy control participants at the mean age for the sargramostim/GM-CSF-treated mild-to-moderate AD participants, 67.8 years (ratio estimate = 1.440, 95% CI: (1.101, 1.885), p=0.0088), whereas the plasma UCH-L1 concentrations of the mild-to-moderate AD participants at the baseline visit prior to GM-CSF/sargramostim treatment are approximately the same as those of the healthy control participants (ratio estimate = 0.967, 95% CI: (0.798, 1.171), p = 0.7257) (**Figure 5C**). UCH-L1 concentrations in MCI were higher than in mild-to-moderate AD at 67.8 years (ratio estimate = 1.490, 95% CI: (1.133, 1.961), p=0.0052). UCH-L1 increased for participants with mild-to-moderate AD by an estimated 2.420% per year of age (95% CI: (0.783%, 4.083%), p=0.0054), while the estimated age-associated increase for MCI is marginally statistically non-significant (estimate = 2.499% per year, 95% CI: (−0.446%, 5.531), p=0.0941), possibly because it is underpowered with the modest sample size. There were no statistically significant differences among the age slopes for healthy controls, mild-to-moderate AD, and MCI for UCH-L1 (interaction test p value = 0.2040). This analysis suggests that during the age-associated progression to mild-to-moderate AD, plasma levels of GFAP and NfL, markers of astrogliosis and axonal damage, respectively, rise with the level of cognitive decline, but UCH-L1 measures of neuronal cell damage are higher early during the MCI phase and are then closer to normal at the mild-to-moderate AD stage, perhaps due to lower neuronal loss, or to lower release of UCH-L1 from neurons.

### Treatment of AD trial participants with sargramostim/GM-CSF reverses the rate of neuronal loss to that of normal controls many decades younger

In addition to identifying potential mechanisms of brain aging, age-associated biomarkers may be used to assess the efficacy of interventions that may slow or even halt the aging process. We have previously discovered GM-CSF/sargramostim as a potential treatment for AD whose likely mechanisms of action may include targeting the aging process in the brain. Specifically, based on early studies showing that RA patients had a reduced risk of developing AD, we hypothesized that this protection might be due to a physiological reaction against RA’s associated inflammation, with the beneficial side effect of reducing the risk of AD, which exhibits brain inflammation. We tested our hypothesis and found that treatment of a mouse model of AD with GM-CSF, an immune system stimulating/modulating cytokine that stimulates the proliferation of immune stem cells in both the bone marrow and the brain and is upregulated in the plasma of RA patients, reduced brain amyloid levels by half, increased brain synaptophysin levels, and restored memory to normal after a few weeks of subcutaneous administration ^35^. This finding was subsequently confirmed in a different mouse model of AD ^36^. Furthermore, we and others have shown that GM-CSF treatment also improves cognition and neuronal function in aged wild-type mice, indicating that the beneficial effects of GM-CSF on the brain are not exclusively targeting AD pathology but also extend to normal aging ^35,37,38^.

Building on this foundation, we recently completed a Phase II, double-blind, randomized, placebo-controlled trial of human recombinant GM-CSF (sargramostim) (250 mcg/m^2^/day subcutaneous injection, five days/week for three weeks) in participants with mild-to-moderate AD ^33^. Treatment with sargramostim led to improved scores in the Mini-Mental State Exam (MMSE) by almost two points (compared to baseline and to placebo) and moved the concentrations of AD-associated plasma biomarkers—amyloid, total Tau—toward normal. This was the first full phase II trial of an intervention that showed improvement, instead of slowing, in AD patients ^33^. Interestingly, the largest change in a measure of AD neuropathology at the end of treatment was in plasma UCH-L1 concentrations, which had decreased in the sargramostim-treated group by 40% compared to baseline (p=0.0017) and by 42% compared to placebo (p=0.0019) ^33^. However, in that study we had no way to assess the true impact of the reduction of plasma UCH-L1 concentrations compared to cognitively normal individuals. Here, we therefore compared the previous results to the age curves for UCH-L1 shown in **Figure 1** to determine how effective sargramostim/GM-CSF treatment was in reducing this measure of neuronal loss with aging.

The plasma concentrations of UCH-L1 in mild-to-moderate AD participants at baseline and at the end of treatment with sargramostim/GM-CSF are plotted with the data from healthy control participants in **Figure 6A** and show that the absolute values of plasma UCH-L1 are greatly reduced after sargramostim/GM-CSF treatment (ratio estimate = 0.497, p=0.0008). Indeed, sargramostim/GM-CSF treatment reduces the concentrations of UCH-L1 in plasma of trial participants to an average level far below those of similarly aged healthy control participants (ratio estimate = 0.502, p=0.0019), and equivalent to that found in healthy control participants six decades younger (**Figure 6A**). For example, a 67.8-year-old mild-to-moderate AD participant treated with sargramostim/GM-CSF would have had a plasma UCH-L1 concentration of 6.47 pg/ml (geometric mean, 95% CI: (4.45, 9.39) pg/ml, not baseline calibrated – sargramostim/GM-CSF baseline only), which is equivalent to that expected of a 5.4-year-old healthy control participant, based on our cross-sectional data. This conclusion required the comparison of the plasma UCH-L1 data from the sargramostim treatment trial to the new data shown in **Figure 1**. The rate of loss of UCH-L1-positive neurons during sargramostim/GM-CSF treatment in the clinical trial is shown in **Figure 6B**: the three-week treatment resulted in a reduction in the rate of UCH-L1 neuron deaths by 2.62 million neuron deaths per year per person (95% CI: (1.53, 3.71) million).

**Figure 6.**
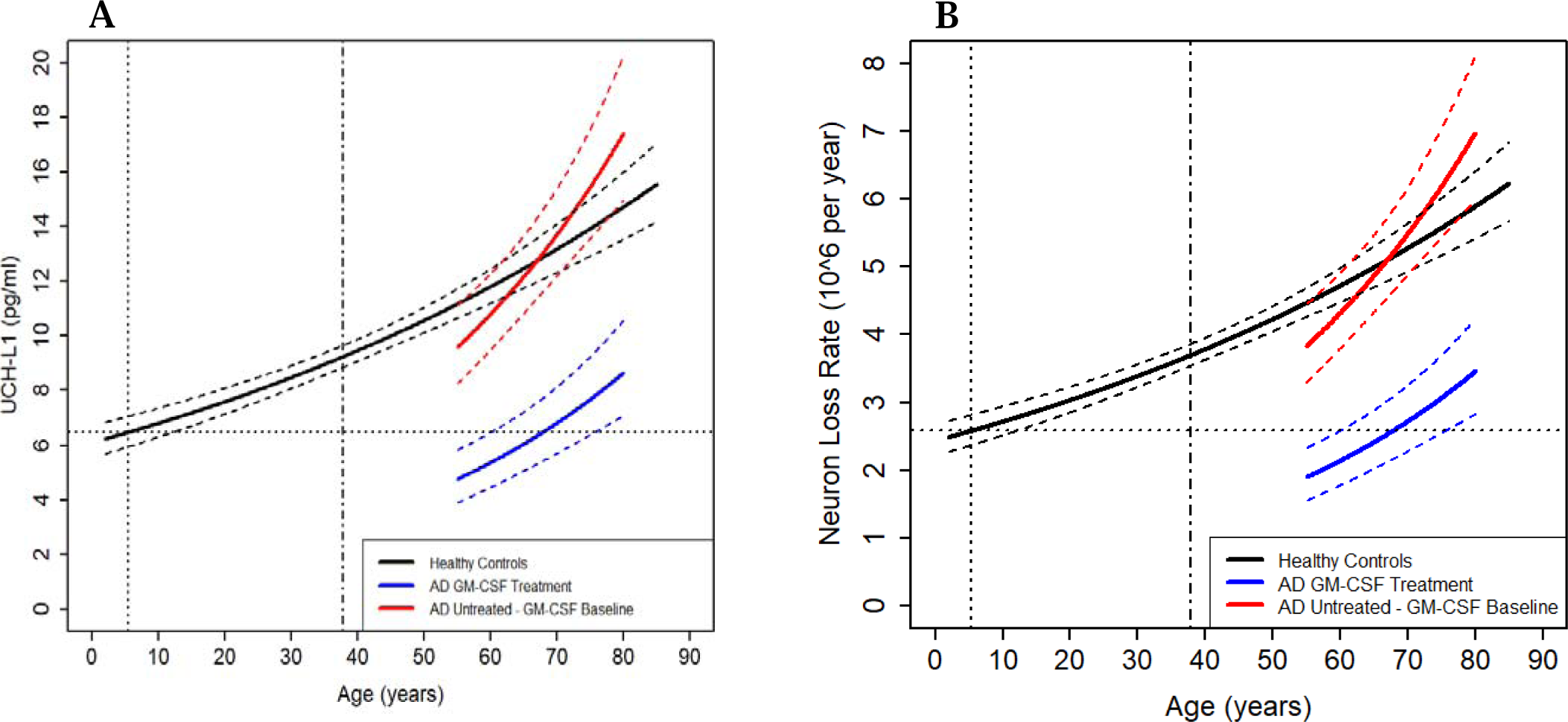
Treatment of participants with mild-to-moderate AD with sargramostim/GM-CSF reduces plasma UCH-L1 concentrations and neuronal loss to levels seen in healthy control participants six and a half decades younger with an average comparable to that of 5.2-year-old healthy control participants. Association plots and point wise standard errors are shown for the participants with mild-to-moderate AD from the Phase II, double-blind, randomized, placebo-controlled trial with recombinant human GM-CSF (the “sargramostim/GM-CSF AD trial”) at baseline before they were treated with sargramostim/GM-CSF (AD Untreated - GM-CSF Baseline, n=18) or after they were treated with sargramostim/GM-CSF (AD GM-CSF Treatment, n=18) together with the correlation curve for healthy control participants from **Figure 1** (**A**). The comparison shows that treatment of participants with mild-to-moderate AD with sargramostim/GM-CSF leads to absolute UCH-L1 values that are greatly reduced compared to their starting (baseline) levels (ratio estimate = 0.497, 95% CI: (0.350, 0.705), p=0.0008). Furthermore, sargramostim/GM-CSF treatment reduces expected plasma UCH-L1 concentrations far below those of similarly aged healthy control participants (ratio estimate = 0.502, 95% CI: (0.341, 0.741), p=0.0019 at a mean age of 67.8 years), and leads to a geometric average plasma UCH-L1 level that sets a 67.8-year-old mild-to-moderate AD participant approximately equal to that expected of a 5.4-year-old healthy control participant (horizontal dotted line). The healthy control age equivalent of the plasma concentrations of UCH-L1 after GM-CSF treatment ranged from around 4 years old for 67-year-old mild-to-moderate AD participants to around 32 years old for 80-year-old mild-to-moderate AD participants. The reduced plasma UCH-L1 levels associated with GM-CSF-treatment of mild-to-moderate AD participants are statistically significantly lower than those of all healthy control participants above approximately age 37.8 (vertical dot-dash line) but are statistically indistinguishable (p>0.05) from healthy control participants younger than age 37.8. The plots calculated for neuron loss are shown in (**B**), which indicate that this three-week treatment resulted in a reduction in the rate of neuron deaths by 2.62 million neuron deaths per year per person (95% CI: (1.53, 3.71) million).

### GM-CSF treatment reverses neuronal apoptosis in hippocampi of AD rats

To investigate the mechanism by which GM-CSF treatment reduces plasma concentrations of UCH-L1 and thus neuronal death, we examined aged TgF344-AD rats (18-20 months of age), a model of AD that shows the complete brain pathology of human AD (amyloid and Tau deposition and neuronal loss) ^39^. TgF344-AD rats were treated with recombinant rat GM-CSF or saline placebo for five weeks, and their brains were assessed by immunohistochemical staining for Caspase-3, a marker of apoptosis that is increased in humans and animal models with AD, and during aging ^40,41^. Data from the treated TgF344-AD rats were also compared to age-matched wild-type (WT) control F344 rats. As shown in **Figure 7**, aged TgF344-AD rats treated with saline exhibited large numbers of Caspase-3-positive cells in the CA1, CA3, and dentate gyrus/hilus regions of the hippocampus compared to WT F344 rats. GM-CSF treatment of aged TgF344-AD rats significantly reduced the elevated number of Caspase-3-positive cells compared to placebo-treated TgF344-AD rats, almost reaching the low number observed in the untreated WT F344 rats. Notably, the vast majority of Caspase-3-positive cells in the TgF344-AD rats were neurons (approximately 95%) based on co-staining for the MAP2 neuronal marker (data not shown). These results indicate that the statistically significant reduction in plasma UCH-L1 in the sargramostim/GM-CSF-treated human AD participants is likely reflecting the biologically significant prevention of AD-associated neuronal apoptosis.

**Figure 7.**
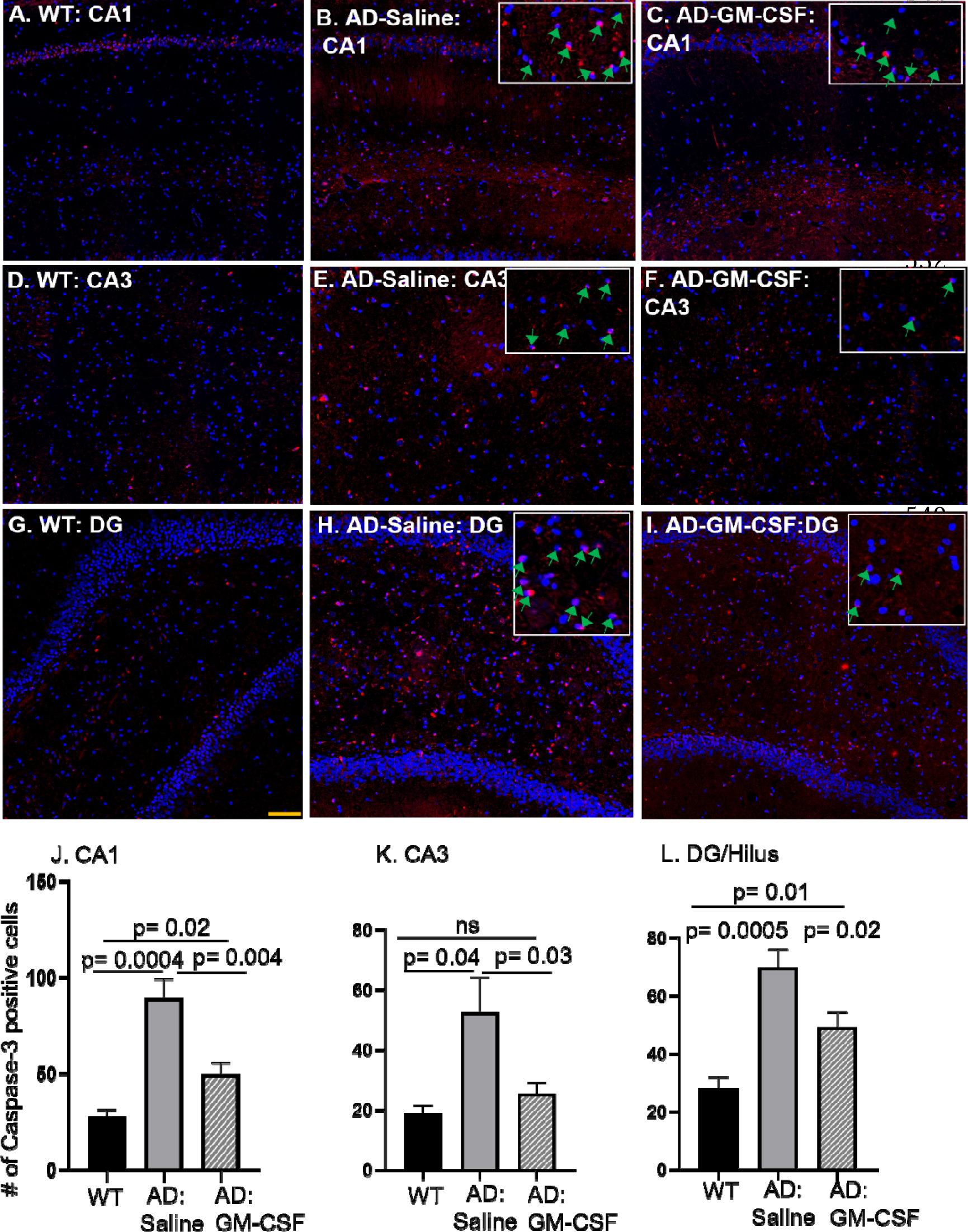
Treatment with GM-CSF reduces neurodegeneration and neuronal cell death in a rat model of AD. Aged male TgF344-AD rats (18-20 months), a model of AD that overexpresses human Aβ peptide ^39^, were treated with GM-CSF or placebo for five weeks as described, and the levels of neuronal damage were assessed by immunohistochemical staining for Caspase-3 (red) together with DAPI staining (blue), followed by blinded counting of Caspase-3-positive cells. Arrowheads in the inserts indicate Caspase-3-positive cells. The numbers of Caspase-3-positive cells were determined in the CA1 (**A-C**), the CA3 (**D-F**), and the dentate gyrus/hilus (**G-I**) regions of the hippocampus in age-matched F344 male wild-type (WT) rats (A,D,G), TgF344-AD rats injected with saline at the end of treatment (B,E,H), and TgF344-AD rats treated with GM-CSF at the end of treatment (C,F,I). Quantitative analyses showed a significantly higher number of Caspase-3-positive cells in all three regions of TgF344-AD rats injected with saline compared to age-matched WT rats (p values as indicated), which decreased significantly with GM-CSF treatment (p values as indicated; J-L). For each bar, data are represented as mean +/-SEM for separate groups of mice. Statistical significance was determined by the unpaired Student’s *t*-test for comparison between groups. Scale bar: 100 μm (20X magnification). All experiments were repeated 2-6 times with similar results (n=5 for WT rats, n=7 for Tg344-AD rats injected with saline, and n=7 for Tg344-AD rats treated with GM-CSF). Notably, the vast majority of Caspase-3-positive cells in the analyzed regions in the TgF344-AD rats were neurons (approximately 95%) as determined by co-staining for the MAP2 neuronal marker (data not shown).

## Discussion

The identification of biomarkers of brain aging is an area of active investigation, and our study has important strengths over previous reports. For example, the identification of biomarkers of aging in different organs, including brain, has been reported recently ^20^. However, that study examined a limited age spectrum and thus does not give insights into when brain aging begins, as we have found. Furthermore, the ‘cognition brain’ markers examined in that study related more to neuronal function than to neuronal loss or damage per se. Most importantly, previous studies have not shown an **exponential** age-associated elevation in markers of brain damage across the life span that our data show.

An exponential increase in any product of a biological process implies the existence of at least one positive feedback loop that accelerates the process. Our finding of exponential rises in the cross-sectionally assessed markers of brain degeneration, first of neuron loss and axon damage, and then of astrogliosis/neuroinflammation with age and their strong correlations with each other evidenced in Figures 1-4 implies the existence of such a positive feedback loop in the process of brain aging. Specifically, the rise in neuron loss and axon damage evident in plasma from early childhood is likely to induce gliosis/inflammation to phagocytose the resulting debris, which would initiate the ‘inflammaging’ cascade, resulting in a vicious cycle of more neuronal damage and death and more inflammation.

Notably, the positive feedback loop involving neuronal loss/damage and inflammation in brain aging throughout life that the new data demand includes the same components as the already-established positive feedback loop that comes into play later, in the development of AD. Specifically, we and others have found that gliosis/neuroinflammation in AD increases the expression of cytokines that lead through several steps to increased production of Aβ peptides and their polymerization into neurotoxic oligomers and filaments. The resulting Aβ oligomers, in turn, further increase neuroinflammation through further activation of microglia and astrocytes ^15,16,42–44^. Such positive feedback loops are ideal targets for developing inhibitors of pathogenic pathways. The other key discovery of our investigation here—that a long-approved immune stem cell stimulating drug, GM-CSF/sargramostim, can be repurposed to reverse age-associated neuronal loss in both human research participants and an animal model of AD—illustrates the benefit of blocking a key step in the feedback loop of age-associated brain degeneration.

In sum, our findings allow several novel conclusions to be drawn:

1. The concentrations of two plasma biomarkers of neuronal damage, UCH-L1 (**Figure 1**) and NfL (**Figure 2**), are exponentially higher with increasing age in healthy control participants, indicating that normal brain aging is a process that starts in early childhood, continues throughout life, and becomes behaviorally apparent only later in life, probably as the neuronal damage overcomes functional redundancy and resiliency.
2. The higher levels of neuron-derived UCH-L1 concentrations in plasma with age in healthy control participants imply an exponential increase in the loss of, or severe damage to, neurons (estimated as a loss of roughly several hundred million neurons over a lifespan, or a minimum of 338 million neuron deaths per person in the sex pooled model (**Supplementary Figure 2**) and also provides an additional ‘N’ measure for the AD ‘AT(N)’ assessment tool. Presumably, neurogenesis, which has also been shown to occur throughout life, replaces some of the neurodegeneration ^45^. Neuronal loss may be larger in specific brain regions, such as the hippocampus, which is relatively small and plays a key role in learning and memory.
3. Based on cross-sectional data, the elevations in the concentrations of the plasma biomarkers of neuronal damage, UCH-L1 and NfL, in healthy control participants occur earlier than the increase in astrogliosis (GFAP), which accelerates significantly after age 40 (**Figure 3**), indicating that astrogliosis and its attendant inflammation (‘inflammaging’) are likely to be both a response to, and an accelerator of, the age-related neuronal damage, rather than an initiating cause. Indeed, this acceleration is evidence of a positive feedback loop that explains the exponential feature of the age-associated increase in all three biomarkers and of the age-associated risk of AD.
4. The higher levels in the measures of neuronal damage and reactive astrogliosis that coincide with increasing age in healthy control participants are highly correlated with each other and may underlie the enhanced risk of developing AD and other neurodegenerative diseases with increased age (**Figure 4**).
5. Because plasma concentrations of NfL (**Figure 5A**) and GFAP (**Figure 5B**) are higher in participants with MCI and mild-to-moderate AD compared to healthy control participants, they appear to reflect active AD-associated injury, with NfL levels being higher after the MCI to AD progression. In contrast, plasma concentrations of UCH-L1 were higher in participants with MCI than in healthy control participants or participants with mild-to-moderate AD (**Figure 5C**), which may reflect greater leakage from damaged neuronal cell bodies early in the disease process compared to healthy aging brains.
6. The higher age-associated levels in plasma concentrations of UCH-L1 as a measure of greater neuronal cell loss in females (**Figure 1C and 1D**) should be further explored to clarify whether this explains or contributes to their increased risk for age-associated neurodegenerative diseases such as AD.
7. Although the standard errors of the mean for the concentration versus age curves for all three biomarkers are very tight across the lifespan, there is variance between individuals. Such variance may reflect increased risk or resilience to the development of AD or AACD caused by genetic variants such as *APOE4* alleles ^43^, lifestyle, environment, or other as-yet unidentified factors^10^.
8. Higher plasma concentrations of UCH-L1 in an individual compared to the expected age-associated level of the healthy control participant curve may serve as a predictive biomarker for the future development or onset of MCI (**Figure 5C**).

Traditional treatments for AD (e.g., cholinesterase inhibitors and memantine) provide only marginal short-term benefits by improving acetylcholine neuronal transmission or modulating excitotoxicity, but they do not modify or target the disease mechanism. The recently FDA-approved anti-Aβ/amyloid monoclonal antibodies, aducanumab, lecanemab, and donanemab remove amyloid deposits effectively in clinical trials, but they may only slow progressive cognitive decline and can lead to amyloid-related imaging abnormalities (ARIAs; brain edema and micro-hemorrhages) and have been linked with smaller brain volumes ^46,47^. Because most current approaches to AD therapy target AD pathologies in symptomatic adults, none are designed or expected to address the contribution of aging to the development of AD.

In contrast to other potential treatments for AD, sargramostim/GM-CSF is also beneficial in normal AACD mouse models and for many neurological injuries and diseases without associated AD pathology, for example in animal models of stroke, TBI, and Parkinson’s disease ^48–51^, in humans with chemobrain ^52^, and in a preliminary clinical trial in participants with Parkinson’s disease ^53^. Its broad therapeutic applicability may be related to the ability of GM-CSF to cross the blood-brain barrier, to be both neuroprotective and anti-apoptotic, to stimulate arteriogenesis and blood flow, to promote axon preservation/regeneration and neuronal plasticity, and to induce the proliferation of neural stem cells ^51,54–58^ (for more references and discussion see ^16^). The finding that sargramostim/GM-CSF treatment in participants with mild-to-moderate AD in the completed Phase II trial led to reduced plasma concentrations of UCH-L1, but not of NfL or GFAP, may be due to the very short treatment period of three weeks. In TBI, plasma UCH-L1 levels increase very rapidly, which dynamically reflects neuronal damage, and then return quickly to baseline after the insult ends, whereas NfL and GFAP are long-lived biomarkers that increase more slowly but persist with longer half-lives and would thus be expected to take longer to show a decrease in response to AD interventions such as sargramostim/GM-CSF ^59^. Our current NIH-funded trial of longer-term treatment (six months) with sargramostim/GM-CSF in participants with mild-to-moderate AD (NCT04902703) may reveal a reduction in plasma concentrations of NfL and GFAP later during the treatment period.

The finding that three weeks of sargramostim/GM-CSF treatment of participants with mild-to-moderate AD in a Phase II, double-blind, randomized, placebo-controlled trial led to improved performance on a cognitive measure (almost 2 points on MMSE) and changes in plasma levels of markers of AD neuropathology (Aβ 40, total Tau, and UCH-L1) ^33^, together with the novel findings reported here, allow the following additional and novel conclusions to be drawn:

9. Sargramostim/GM-CSF treatment of participants with mild-to-moderate AD and rats that model AD effectively halts neuronal cell loss assessed by the plasma concentration of UCH-L1 (**Figure 6**) or Caspase-3/apoptosis (**Figure 7**), respectively.
10. Because a reduction in plasma UCH-L1 was a measure of successful treatment of AD participants with sargramostim/GM-CSF (^33^ and **Figure 6**), UCH-L1 measures may be a sensitive biomarker for testing the efficacy of other AD treatments and therapeutic changes in lifestyle.

Our finding that GM-CSF treatment significantly reduced the elevated apoptosis in the CA1, CA3, and dentate gyrus/hilus regions of the hippocampus in the TgF344-AD rat model of AD to levels that were close to those of age-matched WT control rats (**Figure 7**) suggest that the ability of GM-CSF to improve cognition as measured by MMSE, to partially normalize the concentrations of amyloid and tau plasma biomarkers, and to greatly reduce the plasma concentrations of the biomarker of neurodegeneration, UCH-L1, in our previously published Phase II clinical trial of participants with mild-to-moderate AD ^33^ is likely due to a reduction in the number of apoptotic cells in the brain. This reduction in apoptosis is likely due in part to suppression of apoptosis itself, which GM-CSF has been shown to effect, which would thus increase total levels of UCH-L1 and its synaptic function in the larger number of remaining neurons ^31,56,57^. Furthermore, we and others have found that apoptotic/damaged brain neurons in neurodegenerative diseases, including AD, are often aneuploid, which would lead to substantial changes in gene expression and defects in cellular functions, and that aneuploidy also leads to apoptosis (for review see ^60^). Indeed, cells naturally undergoing senescence accumulate with increased age, and their removal with ‘senolytic’ molecules can reverse some features of aging, including improving cognition in animal models of AD ^42,61–63^. Similarly, through its ability to stimulate the production and activity of innate immune phagocytes (i.e., microglia) in the brain, GM-CSF treatment may enhance the removal of damaged, apoptotic, senescent neurons, just as it caused the rapid reduction of neurotoxic amyloid deposits in the brains of GM-CSF-treated AD mice ^35^ and AD rats (manuscript in preparation), thus allowing the remaining neurons to function more effectively. In sum, our mechanistic studies in TgF344-AD rats are consistent with the conclusion that:

11. GM-CSF treatment has an anti-apoptotic function and possibly a senolytic function that reduces the number of abnormal neurons and thus reverses neuronal defects and cognitive decline due to neurodegenerative disease or aging.

In participants with mild-to-moderate AD, the benefits of GM-CSF treatment in improving cognition and reducing plasma markers of neuropathology, including plasma concentrations of UCH-L1, is evidently temporary. The disease is ongoing and all markers of neuropathology return to almost pre-treatment levels by 45 days after sargramostim/GM-CSF treatment is halted; yet there is still a statistically significant benefit to cognition as measured by MMSE at 45 days post-treatment compared to the placebo ^33^. It remains to be determined whether treatment of healthy aged individuals with sargramostim/GM-CSF will reduce age-associated neuronal damage and reverse AACD only with continuous application or whether it may similarly confer a more long-lived benefit after treatment is halted.

Limitations:

a. Patients with MCI and dementia due to AD were identified by clinical diagnosis, not confirmed with CSF markers or PET. There were no cutoffs used. Thus, we could not categorize participants into A/T/N positivity. Although plasma AD biomarkers are increasingly used, future studies to confirm our findings would benefit from an analysis of CSF samples or from the use of mass spectrometry to analyze plasma biomarkers.
b. Pooled data were obtained from cross-sectional/baseline assessments from different studies with different inclusion/exclusion criteria, which is also a strength. Caution is warranted in interpreting longitudinal *change* in outcomes, as we cannot rule out cohort effects and resulting bias. Future studies should assess these biomarkers longitudinally across the lifespan, and/or pool studies with harmonized recruitment/enrollment techniques to confirm the results.
c. High CVs in UCHL-1 measures may impact the clinical application of these findings, with cutoffs.

## Methods

### Experimental Design

#### Participants

All human subjects research was approved by the Colorado Multiple Institutional Review Board (COMIRB). Specifically, we analyzed healthy control plasma samples from three different studies. Healthy control participants (n=317) who were part of the Crnic Institute Human Trisome Project (HTP, n=103; age range: 2-61 years; 54% female) (COMIRB #15-2170; NCT0284108; Dr. Espinosa), the University of Colorado Alzheimer’s and Cognition Center (CUACC) Bio-AD longitudinal observational study (n=69; age range: 53-83; 70% female) (COMIRB #15-1774; Dr. Bettcher), or the multiple sclerosis (MS) healthy controls biomarker study (termed Nair) (n=145; age range: 16-86; 64% female; 3 participants lacked usable UCH-L1 data) (COMIRB #21-3703; Dr. Nair). See demographics in **Supplementary Table 1**. The HTP is focused on studying biomarkers and clinical features of people with Down syndrome (DS) and includes typical control participants without DS ^64^, and the CUACC Bio-AD study is focused on studying the effect of inflammation on the development of AD ^17^. The Nair MS biomarker study is investigating biomarkers associated with MS and includes healthy control participants. Together, these three healthy control cohorts span ages 2-85.

Using three community-dwelling healthy control cohorts that are diverse and heterogenous with a wide range of ages adds confidence to the cross-sectional measures of the plasma biomarkers. If different populations had different levels and slopes of a marker with age, then pooling them would be averaging them and would cause a deviation away from linearity across the full age range, with parametric linear fit trying to smooth it out. However, there is no appreciable deviation from linearity for UCH-L1 or NfL when examined with the spline fits ^65^ (**Supplementary Figure 1**). GFAP measures across age are more complex, increasing exponentially only after age 40.

Participants assessed as having mild cognitive impairment (MCI) due to AD (n=32) were part of the CUACC Bio-AD longitudinal observational study and were diagnosed based on an interdisciplinary consensus conference with review of cognitive testing, neurological examination, clinical dementia rating scale (CDR), and brain MRI (COMIRB# 15-1774; Dr Bettcher). Participants with mild-to-moderate AD were from our published Phase II, double-blind, randomized, placebo-controlled trial ^33^ with recombinant human GM-CSF (the “sargramostim/GM-CSF AD trial”) (COMIRB # 12-1273; NCT 01409915). We include available plasma samples taken at the baseline visit prior to treatment with sargramostim/GM-CSF or placebo (n=36), as well as available plasma samples taken at the end of three weeks of treatment with either GM-CSF (n=18) or placebo (n=18). Some plasma samples were unavailable for measurements of NfL and GFAP concentrations.

#### Measurement of plasma biomarker concentrations

Concentrations of UCH-L1, GFAP, and NfL in plasma samples were assessed in healthy control participants and in participants with MCI due to AD using published methods ^17,33^ and the Quanterix single molecule array, or SIMOA^®^, SR-X Analyzer system and the Neuro-4-Plex B kits. Concentrations of UCH-L1, GFAP, and NfL in the samples from mild-to-moderate AD participants in the sargramostim/GM-CSF AD trial were determined in our previously published manuscript using the same methods ^33^.

#### Rat AD model

Animal research was approved by the University of Colorado Institutional Animal Care and Use Committee (IACUC#00878). The TgF344-AD rat was developed as a model of AD by inserting transgenes that express the Swedish mutant human *APP* (*APPsw*) and mutant human presenilin 1 (*PSEN1 delta E9*) genes that cause familial AD ^39^. As a result, and due to the fact that the rat MAPT (Tau) gene resembles the human version, the TgF344-AD rats overexpress human Aβ peptide and develop the full complement of human AD brain pathology: amyloid deposits, p-Tau-positive neurofibrillary tangles, and neuronal loss. Untreated age-matched wild-type (WT) F344 rats were used as controls. The 18-to 20-month-old TgF344-AD male rats were injected subcutaneously with GM-CSF (83.3 mg/kg/day; 5 days/week; n=7) or with saline (200 ml/day; 5 days/week; n=7) for 24 injections total over 32 days. On day 32, the rats were anesthetized with sodium pentobarbital, perfused intracardially with PBS for 5-7 min, and the brains were removed rapidly. The right hemisphere was immersed in freshly prepared 4% paraformaldehyde (PFA) in PBS for 24 h at 4 ^◦^C. After fixing with PFA, 4 mm-thick paraffin-embedded hippocampal brain sections were mounted on glass slides and processed for immunohistochemistry for the apoptosis marker Caspase-3 (Cell signaling Technology; Cat# 9662) and stained with DAPI. A detailed protocol for immunohistochemistry and imaging is described in ^38^. We visually counted the numbers of Caspase-3-positive cells in the CA1, CA3, and dentate gyrus/Hilus regions of the hippocampus (blinded as to genotype and treatment).

#### Neuronal death calculation

UCH-L1 is primarily a neuronal protein that is released from damaged neurons in the brain, enters the CSF, and is ultimately released into the blood. Therefore, for each research participant, it is possible to calculate the number of neurons that have released UCH-L1 in a given time period based on the steady state concentration of plasma UCH-L1 (‘C’ in pg/ml from the individual participant data of **Figure 1**), the half-life of UCH-L1 (estimated to be approximately 8.5 hours ^66,67^), the volume of plasma in a typical adult person (3,500 ml), and the amount of UCH-L1 per neuron (2.5% of total protein (250 pg) ^25,26^ = 6.25 pg). Specifically, for each participant, the approximate number of neurons lost per year can be calculated using the following formula:

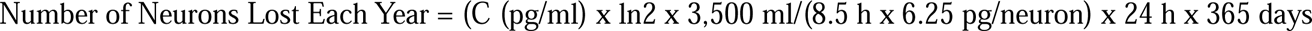

### Statistical Analysis

Data were analyzed using mixed model regression, with unstructured error covariance on repeated measures, for the effects of biomarkers, disease status, and treatment on the logarithmic transforms of the plasma biomarkers of UCH-L1, GFAP, and NfL. Although data from healthy control participants and participants with MCI due to AD were cross-sectional, the framework of mixed model regression could still be adapted, as regression with independent data is merely a simplified version of regression with correlated data. Healthy control, MCI due to AD, mild-to-moderate AD at baseline, mild-to-moderate AD treated with placebo/saline, and mild-to-moderate AD treated with sargramostim/GM-CSF were allowed different covariance matrices. Because the biomarker data were measured using 2 or 3 replicates, all of the observations were used, instead of averaging replicates and weighting. The age effects were modeled as log-linear, with separate age slopes and intercepts for healthy control, MCI due to AD, and mild-to-moderate AD. The intercept for mild-to-moderate AD depended on the treatment x study time, but a common age slope was assumed for all mild-to-moderate AD participants, and independent of treatment effects. The log linearity assumption was checked graphically, and by comparing to spline fits ^68^. Interactions with sex were also considered. Linear combinations of model parameters were estimated, along with 95% confidence intervals, back transformed, and tested. Histograms of the Coefficient of Variation among replicates found some biomarker measures obtained with the SIMOA^®^, SR-X Analyzer system, particularly UCH-L1, for example, in analyses of the effects of TBI, can have Coefficients of Variance much higher than 20%, indicating that the variance is an inherent feature of the measure and not due, for example, to unreliable outliers. For example, a histogram of the UCH-L1 coefficient of variance of the healthy controls is shown below:

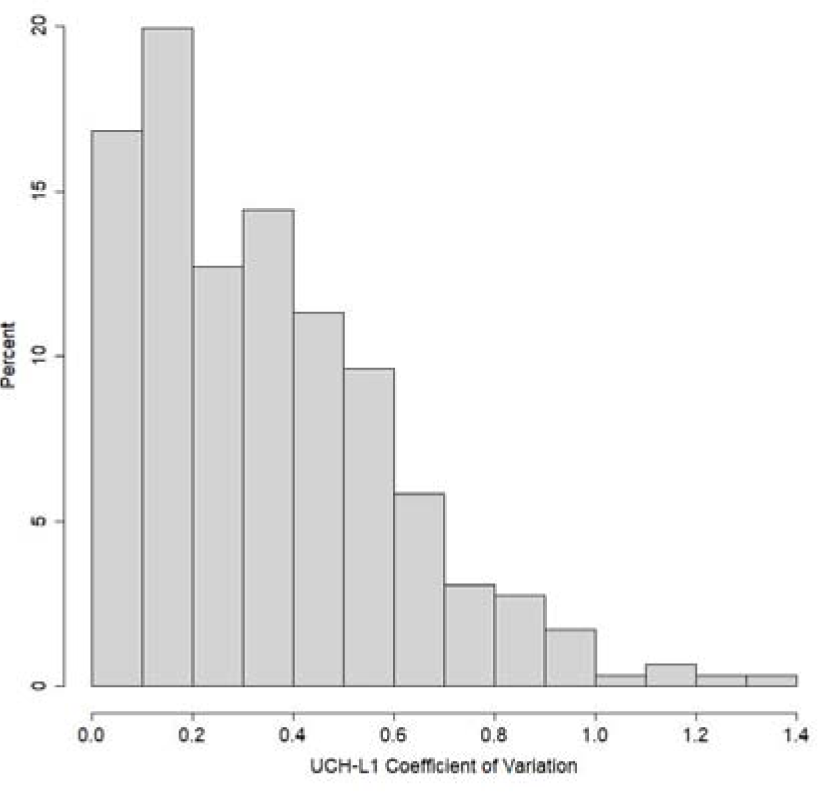

There are known limitations to using biomarkers with high CVs in analyses, with most studies utilizing less than 20% cutoffs at the higher end. The primary concern is that if analysis of the same sample twice yields markedly different values, there are understandable concerns for reliability of measurement (and clinical applicability). Variability of such measures limits their utility for assessing individuals from a small number of replicates, but the laws of large numbers still allow us to estimate the average with a medium to large sample. For cross-sectional comparisons between large cohorts, the average or log average of individual participant’s replicate measures is often used because measures of UCH-L1 often show high variance ^27,66,67,69^. UCH-L1 levels were FDA approved in 2018 as part of a measure of brain damage after TBI ^70^. Because some of our cohorts had two replicates for each participant (healthy controls and participants with MCI due to AD from the Bio-AD) and some (healthy controls from the HTP, GM-CSF trial participants with mild-to-moderate AD, and healthy controls from the MS study) had mostly three replicates for each participant, and some of the biomarkers, especially UCH-L1, have high variability among replicates, we decided in the modeling to use each replicate as a separate measure in the main analysis, rather than using the averages of the individual participants’ replicate measures. The variation among replicates was modeled by introducing an additional noise term into the model. A common variance for the replicate noise term was assumed across all treatments because of software option limitations. Model predicted values of the response, along with pointwise standard errors, were plotted. All data calculations are provided in the **Supplementary Tables 2-4**. As a test of confidence in the results, the averages of the participants’ replicate measures for the biomarkers were also modelled, weighted by the number of replicates, and the conclusions of age-associated exponential increases in the three biomarkers, UCH-L1, NfL, and GFAP, and the effect of GM-CSF treatment on reducing UCH-L1 levels were obtained. The overall results and conclusions were the same. Univariate statistical significance was set at alpha = 0.05, two-sided, for all tests unless otherwise stated. Statistics were computed using SAS 9.4 and R 4.1.3. There are remaining limitations to using this approach in terms of both within-site and cross-site replication of findings, and we acknowledge that it is challenging to move a biomarker into clinical practice for AD with high CVs, although it has been done for UCH-L1 in TBI.

## Supporting information

demographics

## Data Availability

All data produced in the present study are available upon reasonable request to the authors

## Acknowledgements

National Institutes of Health grant R01AG071151 (HP)

National Institutes of Health grant R01AI50305, (JE)

The State of Colorado (HP)

Alzheimer’s Association Part the Cloud grant PTC C-16-422172 (HP)

The Global Down Syndrome Foundation (HP, JME)

The Anna and John J. Sie Foundation (JME)

The University of Colorado Human Immunology and Immunotherapy Initiative (HI3).

## Author contributions

Conceptualization: HP, SHS, NE, HJC

Methodology: SHS, CC, MMA, KN, BMB

Investigation: SHS, CC, MMA, KN, PA, MDG, BMB, JME

Supervision: HP, JME

Writing—original draft: HP

Writing—review & editing: HJC, SHS, CC, MMA, BMB, HP, TDB

## Declaration of generative AI and AI-assisted technologies

None used.

## Data and materials availability

All data are available in the main text or the supplementary materials.

