## supplementary material for "Neuron loss in the brain starts in childhood, increases exponentially with age and is halted by GM-CSF treatment in Alzheimer’s disease"

A

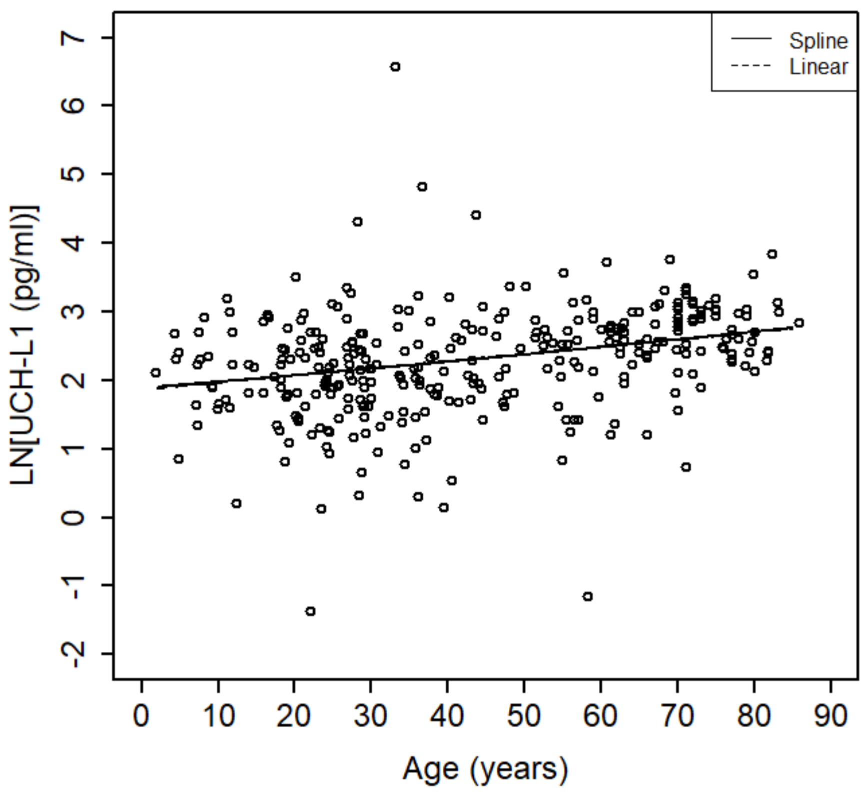
Spline
Linear
LN[UCH-L1 (pg/ml)]
Age (years)

B

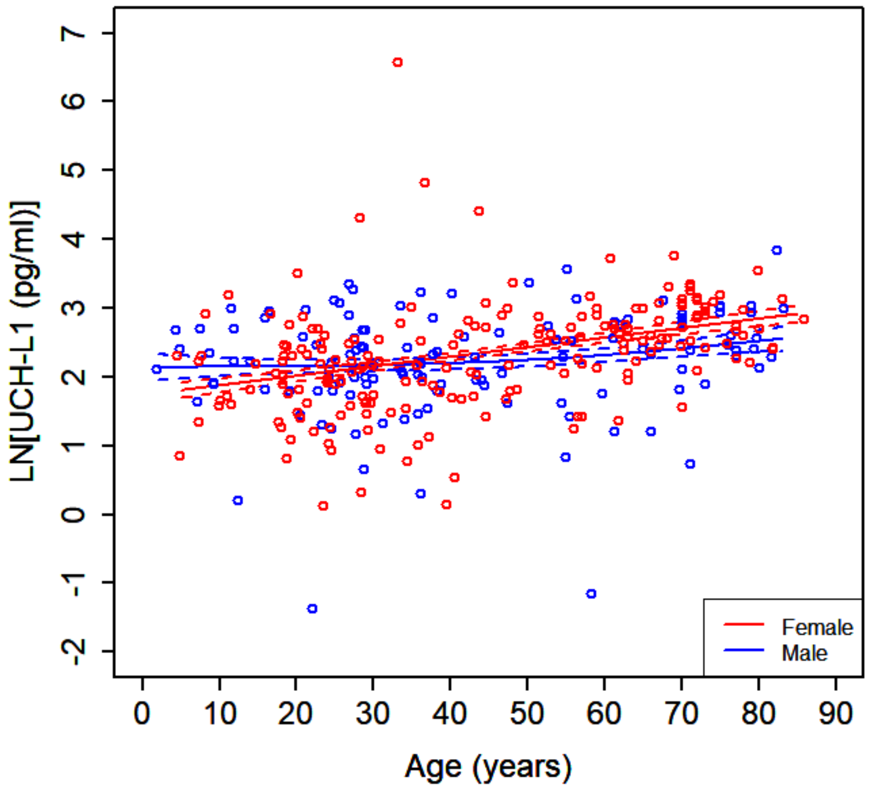
Female
Male
LN[UCH-L1 (pg/ml)]
Age (years)

C

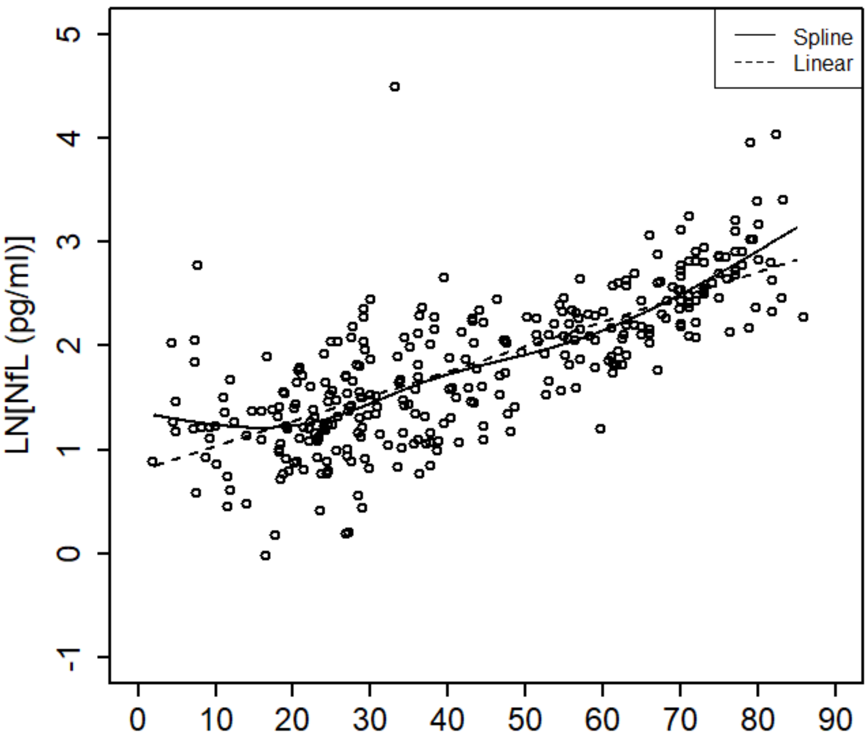

Spline
Linear
LN[NfL (pg/ml)]
5
4
3
2
1
0
-1
0
10
20
30
40
50
60
70
80
90


D

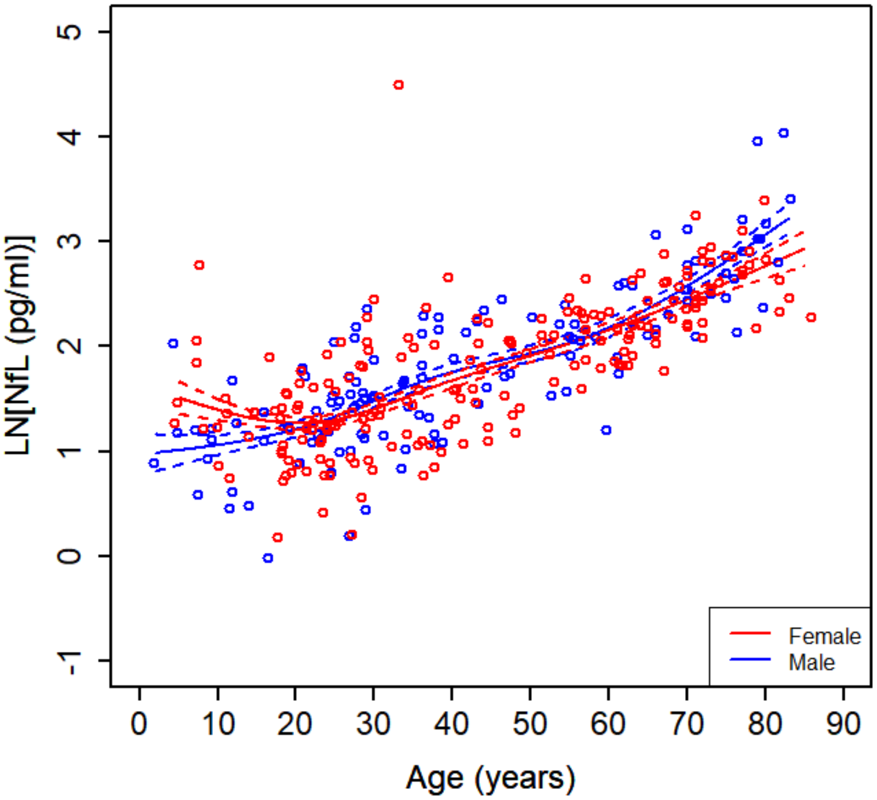

Female
Male
LN[NfL (pg/ml)]
5
4
3
2
1
0
-1
0
10
20
30
40
50
60
70
80
90
Age (years)

**Supplementary Figure 1.** Comparing log linear fit to splines for biomarker association with age and sex in healthy controls (replicates averaged) for:
**UCH-L1 (A):** Deviance test for comparing spline model to the null hypothesis of linear: p value = 0.199. There is no statistical evidence against linearity even with a large sample. **(B):** Spine plot by sex showing no significant deviation from linearity in either males or females.
**NfL (C):** Deviance test for comparing spline model to the null hypothesis of linear: p value = 0.000125. Evidence against perfect linearity with a large sample, with the graph suggesting that the rate of increase accerates somewhat with older age, but linear is still a reasonable approximation. **(D):** Spine plot by sex showing similar deviation from linearity in males and females.

**Supplementary Figure 2**. The effects of age and sex on neuronal cell death/damage (from age 5) as assessed by plasma UCH-L1 concentrations.

Because UCH-L1 is derived from damaged brain neurons and released into the CSF and ultimately into the plasma, its concentration in plasma offers a measure of the ongoing neuronal cell death in each individual. The plasma concentration of UCH-L1 can be considered the steady state of a pharmacokinetic condition in which UCH-L1 protein is released into the plasma from neuronal cell death and removed at a known half-life that can be determined by how quickly the UCH-L1 concentration drops with time after an acute TBI injury, which we calculated using a formula shown in the **Methods**. The supplementary Figure 2 shows a cross-sectional plot of the estimated cumulative number of neuron deaths per person in healthy controls versus their age starting at age 5, which results in an exponential increase in neuronal cell death with age, as expected from the increase in UCH-L1 plasma concentrations with age in **Figure 1C.** Because this analysis assumes that all UCH-L1 is released from each dying neuron as a soluble protein and available for assessment in the plasma, the calculation provides a significant underestimate.
The relationship between neuron loss and age is increasing exponential, which indicates that it compounds in a positive feedback loop. The cumulative curve for males is initially higher at young ages. The female cumulative curve with its higher rate of exponential increase surpasses the male curve at older ages.
Between ages 2 and 85, a minimum estimated 338 million neuron deaths occur per person (95% CI: (311, 369) million) in the sex pooled model. Between ages 5 and 83, the overlap for the samples in females and males, an estimated 335 million neuron deaths occur in females (95% CI: (298,377) million), and an estimated 295 million neuron deaths occur in male (95% CI: (268, 324) million). This analysis does not account for differences in neuronal loss (possibly larger) in specific brain regions, such as the hippocampus, which is relatively small and plays a key role in learning and memory.

**Data S1.**

Demographics of all participants

**Data S2**

Plasma biomarker measures of all participants
