## Supplementary material for "Neuron loss in the brain starts in childhood, increases exponentially with age and is halted by GM-CSF treatment in Alzheimer’s disease": demographics

### The FREQ Procedure

Frequency
Row Pct

| Table 1 of Treatment by sex<br>Controlling for data_set=Leukine Study | sex | | |
|---|---|---|---|
| Treatment | Female | Male | Total |
| External AD | 0<br>. | 0<br>. | 0 |
| External Controls | 0<br>. | 0<br>. | 0 |
| External MCI | 0<br>. | 0<br>. | 0 |
| External T21 | 0<br>. | 0<br>. | 0 |
| Leukine | 12<br>60.00 | 8<br>40.00 | 20 |
| Placebo | 11<br>55.00 | 9<br>45.00 | 20 |
| Total | 23 | 17 | 40 |

Frequency
Row Pct

| Table 2 of Treatment by sex<br>Controlling for data_set=Bio AD | sex | | |
|---|---|---|---|
| Treatment | Female | Male | Total |
| External AD | 6<br>46.15 | 7<br>53.85 | 13 |
| External Controls | 48<br>69.57 | 21<br>30.43 | 69 |
| External MCI | 16<br>50.00 | 16<br>50.00 | 32 |
| External T21 | 0<br>. | 0<br>. | 0 |
| Leukine | 0<br>. | 0<br>. | 0 |
| Placebo | 0<br>. | 0<br>. | 0 |
| Total | 70 | 44 | 114 |

Frequency
Row Pct

| Table 3 of Treatment by sex<br>Controlling for data_set=HTP | sex | | |
|---|---|---|---|
| Treatment | Female | Male | Total |
| External AD | 0<br>. | 0<br>. | 0 |
| External Controls | 56<br>54.37 | 47<br>45.63 | 103 |
| External MCI | 0<br>. | 0<br>. | 0 |
| External T21 | 150<br>47.47 | 166<br>52.53 | 316 |

#### The FREQ Procedure

| Frequency<br>Row Pct |
|---|

Table 3 of Treatment by sex

Controlling for data_set=HTP

| Treatment | sex Female | sex Male | Total |
|---|---|---|---|
| Leukine | 0<br>. | 0<br>. | 0 |
| Placebo | 0<br>. | 0<br>. | 0 |
| Total | 206 | 213 | 419 |

| Frequency<br>Row Pct |
|---|

Table 4 of Treatment by sex

Controlling for data_set=Nair

| Treatment | sex Female | sex Male | Total |
|---|---|---|---|
| External AD | 0<br>. | 0<br>. | 0 |
| External Controls | 93<br>64.14 | 52<br>35.86 | 145 |
| External MCI | 0<br>. | 0<br>. | 0 |
| External T21 | 0<br>. | 0<br>. | 0 |
| Leukine | 0<br>. | 0<br>. | 0 |
| Placebo | 0<br>. | 0<br>. | 0 |
| Total | 93 | 52 | 145 |

#### The MEANS Procedure

| Analysis Variable : Age | | | | | | | | | | |
|---|---|---|---|---|---|---|---|---|---|---|
| data_set | Treatment | N Obs | N | Mean | Std Dev | Minimum | Lower Quartile | Median | Upper Quartile | Maximum |
| Leukine Study | Leukine | 20 | 20 | 66.9500000 | 6.4845971 | 56.0000000 | 62.0000000 | 67.5000000 | 71.0000000 | 80.0000000 |
| | Placebo | 20 | 20 | 70.1500000 | 6.4176976 | 55.0000000 | 66.0000000 | 72.5000000 | 74.5000000 | 78.0000000 |
| Bio AD | External AD | 13 | 13 | 66.9230769 | 9.0596315 | 54.0000000 | 57.0000000 | 68.0000000 | 73.0000000 | 81.0000000 |
| | External Controls | 69 | 69 | 69.5072464 | 6.3630902 | 53.0000000 | 65.0000000 | 70.0000000 | 73.0000000 | 83.0000000 |
| | External MCI | 32 | 32 | 73.6562500 | 5.8287905 | 63.0000000 | 70.0000000 | 73.5000000 | 78.5000000 | 87.0000000 |
| HTP | External Controls | 103 | 103 | 28.1242984 | 15.2107619 | 1.7998047 | 14.5996094 | 27.6992188 | 38.6953125 | 61.2968750 |
| | External T21 | 316 | 316 | 23.1516932 | 12.2932819 | 0.9772949 | 13.9492188 | 22.7968750 | 31.7656250 | 57.5937500 |
| Nair | External Controls | 145 | 145 | 40.8688578 | 18.9390208 | 16.3984375 | 24.5000000 | 35.7968750 | 54.8984375 | 85.8906250 |
